# Tackling inequalities in preconception health and care: barriers, facilitators and recommendations for action from the 2023 UK Preconception EMCR Network conference

**DOI:** 10.1101/2024.02.13.24302690

**Authors:** Danielle Schoenaker, Jennifer Hall, Catherine Stewart, Stephanie J. Hanley, Emma H. Cassinelli, Madeleine Benton, Alexandra Azzari Wynn-Jones, Mehar Chawla, Sinéad Currie, 2023 UK Preconception EMCR Network conference workshop participants

## Abstract

Reducing inequalities in preconception health and care is critical to improving the health and life chances of current and future generations. A hybrid workshop was held at the 2023 UK Preconception Early and Mid-Career Researchers (EMCR) Network conference to co-develop recommendations on ways to address inequalities in preconception health and care. The workshop engaged multi-disciplinary professionals across diverse career stages and people with lived experience (total n=69). Interactive discussions explored barriers to achieving optimal preconception health, driving influences of inequalities, and recommendations. The Socio-Ecological Model framed the identified themes, with recommendations structured at interpersonal (e.g. community engagement), institutional (e.g. integration of preconception care within existing services) and environmental/societal levels (e.g. education in schools). The co-developed recommendations provide a framework for addressing inequalities in preconception health, emphasising the importance of a whole-systems approach. Further research and evidence-based interventions are now needed to advance the advocacy and implementation of our recommendations.

## Background

The health, behaviours, and social and economic circumstances of individuals before conception (preconception health) influence their lifelong health, and are key determinants of a successful pregnancy as well as the optimal health and development of any children they may have.^1^ In the UK, most people are not well prepared for pregnancy. Based on the most recent data from the UK, about half of pregnancies are unplanned,^2^ and nine in 10 women enter pregnancy with at least one potentially modifiable risk factor for pregnancy and birth complications, including smoking, obesity, and lack of folic acid supplement use.^3^ These risk factors are common among both women and men across the reproductive years,^3, 4^ including those who are actively planning pregnancy.^5, 6^

Suboptimal pregnancy planning and preconception health disproportionality affect subgroups of the population. In England, for example, women from Black ethnic backgrounds are 1.5 times more likely to live with obesity when entering pregnancy compared with White women (34% vs 23%) and women living in the most compared with least deprived areas are 3-times more likely to smoke around the time of conception (30% vs 10%).^3^ These social and economic inequalities account for a substantial proportion of severe adverse pregnancy outcomes, with 17% of babies born with fetal growth restriction and 24% of stillbirths attributable to ethnicity and socioeconomic deprivation, respectively.^7^ Moreover, the leading causes of maternal and perinatal mortality, including suicide and birth defects,^8, 9^ are influenced by preconception risk factors, such as mental health conditions and lack of folic acid supplement use.^10, 11^ These are in turn more common among Black women and/or those living in the most deprived areas,^3^ which are wider determinants of health that are often intersectional alongside other structural barriers. Improving the health and life chances of current and future generations requires urgent action, particularly to reduce inequalities in preconception health and care.

Optimising preconception health is a priority for the UK government.^12, 13^ However, clear actions to effectively reduce inequalities in preconception health and care in the UK are lacking. An interactive workshop was held at the 2023 UK Preconception Early-and Mid-Career Researchers (EMCR) Network conference to co-develop recommendations on ways to address inequalities in preconception health and care. This paper presents findings from the workshop.

## Methods

### UK Preconception EMCR Network conference

The UK Preconception EMCR Network conference was a 1-day hybrid event held on 16 October 2023 at the University of Birmingham. The event was organised by the UK Preconception EMCR Network, which is a subgroup of the UK Preconception Partnership (https://www.ukpreconceptionpartnership.co.uk/). The conference explored approaches for the development and implementation of interventions through a keynote presentation, and showcased research conducted by students and EMCRs in presentation and panel sessions. These sessions were followed by an interactive workshop titled ‘What is needed to reduce inequalities in preconception health and care?’. The conference was attended by 104 delegates (44 in person and 60 online), who were all invited to participate in the workshop.

### Workshop: What is needed to reduce inequalities in preconception health and care?

The workshop was attended by 69 participants (42 in person and 27 online). Participants were students (26%), early-career (29%), mid-career (17%) or senior professionals (14%), and people with lived experience (i.e. members of the public) (14%). Attendees came from England (75%), Scotland (6%), Wales (4%), Northern Ireland (4%) or another country (11%). Among students and professionals, study or work was mainly focussed on research (66%), clinical practice (24%), policy (6%) or other (e.g. teaching) (4%). Expertise and disciplines were wide-ranging and included (but were not limited to) preconception health and care, maternal and child health, sexual and reproductive health, obstetrics and gynaecology, primary care, public health, epidemiology, behaviour change, and intervention (co-)development.

The agenda for the 75-minute workshop was developed by the conference organising committee (DS, JH, SC, SJH, EHC, MB, AAWJ, MC, SC). DS and JH co-chaired the workshop and delivered a 10-minute presentation on the topic of inequalities in preconception health and care at the start of the workshop. The term ‘inequalities’ was defined as: ‘Differences in health and care across the population (in this case people who may become pregnant or a parent), that are systematic, unfair and avoidable. They are caused by the conditions in which we are born, live, work and grow’.^14^ At the end of the presentation, participants were given three discussion questions:

1. What might prevent people from accessing preconception care and being healthy and well before pregnancy?
2. How does this lead to or increase inequalities in preconception health and care?
3. How can we tackle inequalities in preconception health and care? What are your recommendations and for whom?

Participants were randomly allocated into groups of 5-10 people, both in person (five groups based on colour indication on name tags) and online (three groups based on random allocation into Zoom breakout groups). Groups discussed the three questions over 35-minutes and made notes on flipchart paper or Zoom whiteboards. During the final 30 minutes, one person in each group summarised the key points that were discussed for each question. The presentation and summaries of group discussions were recorded via Zoom.

### Identifying themes and recommendations

After the workshop, notes on each group’s flipchart paper and online whiteboards were transcribed verbatim into a shared word document by DS and SC. CS listened to the workshop recording and added any points which were discussed verbally to the document. DS and SC independently categorised the transcripts into naturally occurring themes (defined as a common, recurring concept with aggregated meaning). These were then discussed, consolidated and refined. CS triangulated the refined themes and found no disagreements. DS and SC generated recommendations based on themes. These were structured using the Socio-Ecological Model (SEM),^15^ which recognises the complex, multifaceted and interrelated influences on health and behaviours specifically for health promotion. It identifies four levels of influence on health: intrapersonal, interpersonal, institutional, and environmental/societal. These levels are applied in this paper using the following definitions:

- Intrapersonal: individual level influences of health and behaviour such as knowledge, motivation, intention, confidence.
- Interpersonal: the influence of other people and groups on health and behaviour such as family and friends, colleagues, community-based organisations.
- Institutional: the influence of community conditions, availability and access to healthcare professionals and services, and recreational facilities.
- Environmental/societal: the political, social, economic, cultural and policy influences on health and behaviour such as nationally published health guidelines, educational curriculum, healthcare budgets.

The themes and recommendations were reviewed and suggested changes provided by other members of the conference organising committee, after which DS and SC discussed the feedback and made minor edits to the recommendations. Subsequently, the workshop findings were circulated via email and reviewed by workshop participants who either agreed or made suggested changes to the themes and recommendations. All feedback was addressed by DS and SC, and minor edits discussed and agreed via email with the conference organising committee.

## Results

Based on the three questions discussed during the workshop, findings are presented on the following topics: 1) barriers to optimal preconception health and care; 2) driving influences of inequalities; and 3) recommendations for research, clinical practice and policy to tackle inequalities in preconception health and care.

### 1) Barriers to optimal preconception health and care

In response to the workshop question ‘What might prevent people from accessing preconception care and being healthy and well before pregnancy?’, a range of barriers were identified. These were grouped into 12 overall barriers, with more detailed notes from the whiteboards and discussions summarised as examples (**Table 1**).

**Table 1.** Barriers to accessing preconception care and being healthy and well before pregnancy.

| Barrier | Examples | Socio-ecological level(s) |
| --- | --- | --- |
| Many pregnancies are not planned |  | Intrapersonal<br>Interpersonal |
| Lack of preconception care services | • Preconception care services are not routinely provided<br>• Lack of awareness of services<br>• Lack of awareness of relevant healthcare professional(s) to talk to<br>• Lack of relevant training and support for healthcare professionals to raise awareness and provide preconception information and care<br>• Lack of time to provide (healthcare professionals) or seek (public) preconception care | Institutional |
| No regular access to healthcare | • Many young people do not have regular contact with healthcare (lack of health seeking behaviour)<br>• National shortage of GPs leading to inadequate access, while alternative (digital) sources of information may not be evidence-based, provide inconsistent messaging, or are not accessible to everyone | Institutional<br>Environmental/societal |
| Lack of holistic healthcare | • Conditions (such as diabetes, epilepsy, mental health conditions) are often treated in silos without consideration of pregnancy intention and preconception care<br>• Lack of continuity of care / clear referral pathways<br>• Lack of a life course approach to reproductive healthcare | Institutional<br>Environmental/societal |
| Lack of knowledge and awareness | • The public are not aware that pregnancy planning and preparation is something important to consider (e.g. many people may not seek advice or care until they have issues with conception)<br>• Lack of education and information for the public (and healthcare professionals) on the importance of preconception health (including teenagers and adults)<br>• Information about pregnancy and parenthood is often focussed on the pregnancy and postpartum periods<br>• Lack of useful terminology and appropriate communication (e.g. the terms 'preconception health' and 'preconception care' are not widely known or understood)<br>• Pregnancy planning and preparation are not addressed as a topic in education, and information is rarely shared through family and friends<br>• Misinformation and myths about preconception interventions exist (e.g. 'folic acid helps fertility' or 'if you have a pre-existing health condition you should not get pregnant') | Intrapersonal<br>Environmental/societal |
| Pregnancy planning and preparation are often seen as a taboo topic or private matter | • Lack of societal awareness and support for preconception care including lack of accessible and inclusive information and education<br>• Social norms around (not) discussing pregnancy planning, preparation and preconception health<br>• Wanting to keep pregnancy intention private | Intrapersonal<br>Environmental/societal |
| Competing priorities of healthcare system | • Major health conditions, such as mental health conditions, are often treated in silos without a holistic approach to healthcare<br>• Focus on current individual health and social priorities, not future or preventative health<br>• Unclear which health professionals are responsible for provision of preconception care (e.g. the delivery of advice) | Institutional<br>Environmental/societal |
| Domestic violence | • Reproductive coercion<br>• Unable to access healthcare | Interpersonal |
| Widespread health inequalities | • Marginalised groups are less likely to access healthcare<br>• Language, cultural and religious barriers<br>• Discrimination related to sexuality and gender roles<br>• One size does not fit all | Intrapersonal<br>Interpersonal<br>Institutional<br>Environmental/ societal |
| Institutional bias | • Bias related to gender, race, ethnicity, social class, sexuality (for example in the workplace, or healthcare professionals' assumptions) | Institutional |
| Structural barriers | • Risky health behaviours are often considered individual responsibility<br>• Cost-of-living crisis and the cost of being healthy<br>• Food insecurity<br>• Housing insecurity | Intrapersonal<br>Institutional<br>Environmental/ societal |
| Lack of self-agency and beliefs | • Lack of confidence and ability to seek information/advice/support<br>• Lack of motivation to optimise health<br>• Negatively framed messages causing blame and stigma<br>• Belief that optimising health before pregnancy is not relevant at the moment (while recognising it may become relevant in the future)<br>• Not relating to information about preconception care | Intrapersonal<br>Interpersonal |

### 2) Driving influences of inequalities

Workshop participants identified how the identified barriers can lead to or increase inequalities in preconception health and care. This included the influence of barriers (e.g. lack of knowledge and access to care) on people’s ability to make informed choices, and on opportunities to seek advice while feeling safe and understood (e.g. influenced by gender, sexuality, language, cultural and religious barriers). Structural barriers such as dealing with the cost-of-living crisis may require people on lower incomes to prioritise immediate day-to-day issues and thereby prevent them from ‘forward thinking’ (e.g. taking the time to plan and prepare for pregnancy). Many barriers (e.g. unplanned pregnancies, lack of appropriate education and information, domestic violence) often co-occur and are more common among people who already face structural barriers (e.g. income, housing, food insecurity). This increases inequalities in preconception care seeking behaviour, and pregnancy planning and preconception health. In addition to barriers leading to inequalities, inequalities may in turn lead to barriers. This may result in a negative cycle, perpetuating the intergenerational transmission of adversity.

### 3) Recommendations

Recommendations to tackle inequalities in preconception health and care were structured using the SEM (**Table 2**). Workshop participants discussed recommendations at interpersonal, institutional and environmental/societal levels. No recommendations were identified at the intrapersonal level, and it was noted that individual-level interventions may increase inequalities especially if they are influenced by structural barriers. Recommendations were relevant to research (e.g. embed stakeholder involvement in the co-development of communication methods, interventions and services), clinical practice (e.g. integrate preconception care within existing services already accessed by the target population) and policy (e.g. provide preconception health and care education in schools).

**Table 2.** Recommendations to tackle inequalities in preconception health and care structured using the Socio-Ecological Model.

| Socio-ecological level | Recommendation topic | Recommendation |
| --- | --- | --- |
| Interpersonal | Community engagement | • Work with and develop community ambassadors for preconception care in marginalised communities (e.g. through community organisations) |
|  | Wide range of information sources and signposting | • Educate and raise awareness among communities and healthcare professionals about preconception health and care |
|  | Communication | • Communicate about preconception health and care using health promotion messages and positive (benefit-based) messages (ultimately from within the community)<br>• Normalise conversations about preconception health and care and reduce shame and stigma (across communities, healthcare, government)<br>• Promote 'shared responsibility' of optimising and reducing inequalities in pregnancy planning and preconception health (individuals, health service, government) |
| Institutional | Community engagement | • Embed stakeholder involvement (including patient and public involvement and engagement) in the co-development of (culturally and socially acceptable) communication methods, interventions and services for preconception care<br>• Work with and develop community ambassadors for preconception care in marginalised communities (e.g. through pharmacies, as well as role models and influencers) |
|  | Integrate/embed preconception care within existing services | • Integrate/embed preconception care within existing services already accessed by the target population (e.g. primary care or postnatal care)<br>• Learn from best practice models in other countries (e.g. explore if approaches to pregnancy intention screening being tested in other countries are acceptable and effective for use in the UK context)<br>• Ensure preconception care is delivered by a diverse workforce (e.g. diverse in terms of gender, ethnicity, cultural background)<br>• Target at-risk groups (e.g. groups with pre-existing conditions or vulnerable status) |
|  | Healthcare professional training | • Increase knowledge and skills in providing preconception care and approaching the topic of pregnancy intention and planning and preconception health across (community-based) healthcare professional domains (e.g. primary healthcare, midwives, etc)<br>• Provide opportunistic preconception care (e.g. across primary care and through postnatal education)<br>• Reduce healthcare professionals' assumptions (e.g. about pregnancy intentions, preconception health knowledge) |
| Environmental/societal | Ensure logistic accessibility of healthcare | • Ensure preconception care is logistically accessible locally (for example through Women's Health Hubs), and supported through low-cost transport, parking and childcare |
|  | Wide range of trustworthy and reliable information sources and signposting | • Provide preconception health and care education in schools (including primary schools and universities)<br>• Raise awareness through wide-ranging signposting about where and how preconception care can be accessed<br>• Develop, disseminate and signpost to a range of resources about pregnancy planning, contraception and preconception health, including through social media campaigns, as well as digital and non-digital resources in multiple languages to ensure accessibility |
|  | Whole systems approach | • Ensure political will at the highest possible level<br>• Address structural/societal/political issues to improve access to healthcare and enable community and individual change<br>• Conduct research that highlights the scope of preconception health and care inequalities and structural barriers (rather than a predominant research focus on how to change individuals' behaviours) – and use these research findings to advocate for policy change |
|  | Promote preconception care pathways | • Provide incentives to increase awareness of the importance, and to drive the implementation, of policy/NICE recommendations (e.g. through a QOF indicator) |
|  | Life course approach | • Educate about preconception health behaviours from a young age through education, healthcare and cultural opportunities |

NICE, National Institute for Health and Care Excellence; QOF, Quality and Outcomes Framework

## Discussion

The workshop brought together multi-disciplinary professionals across all career stages as well as people with lived experience to co-develop recommendations on how inequalities in preconception health and care may be addressed. Key barriers to accessing preconception care and being healthy and well before pregnancy were identified, including lack of preconception care services and awareness. These barriers may prevent appropriate support in preparation for pregnancy, and thereby increase inequalities. Recommendations for research, clinical practice and policy were developed at the interpersonal, institutional and environmental/societal levels, with the aim to support advocacy and action to acknowledge and reduce inequalities in preconception health and care.

Many barriers to being healthy and well (e.g. food insecurity, weight stigma), driving influences of inequalities (e.g. inability to make informed choices and adopt healthy behaviours), and recommendations to reduce inequalities (e.g. taking a whole-systems and life course approach) are relevant not only to preconception health but to health inequalities in general. Health inequalities are driven by a complex range of factors, and therefore require high-level collaborative action across sectors (e.g. health care, food, tobacco and alcohol industry) and government departments (e.g. Health and Social Care, Education, Environment, Food and Rural Affairs, Levelling Up, Housing and Communities, and Transport). Our co-developed recommendations also suggest that local approaches, such as working with community ambassadors in marginalised communities, are important to ensure interventions and initiatives are relevant and tailored to those who need it most.

This is in line with UK government policies such as the Levelling Up White Paper and Core20PLUS5 approach, which aim to reduce inequalities by improving population-level health and care as well as targeting the health of the most disadvantaged groups and areas.^16, 17^ If implemented successfully, these strategies will not only benefit the general population, but will help improve the health, wellbeing and wider circumstances of people across the reproductive years with far reaching benefits for future generations.

Our workshop findings also identified barriers and drivers of inequalities specific to accessing care, and being healthy and well during the months/years before pregnancy and among people of reproductive age generally. Most notable were barriers related to unplanned pregnancies, lack of preconception care services, and lack of appropriate education and information to raise awareness among the public, healthcare professionals and policy makers that pregnancy planning and preparation is something important to consider. Similar barriers are commonly reported in the literature based on a systematic review of 28 studies from the USA, UK, Netherlands, Canada and Australia.^18^ Moreover, lack of motivation to optimise health for a possible pregnancy and child in the future, and the belief that optimising health before pregnancy is not relevant (yet), were also identified as barriers during our workshop and may be influenced by structural barriers. Barker et al. defined four preconception action phases that individuals move through in relation to their goals to become a parent (i.e. children and adolescents; adults with: no immediate intention to become pregnant; intention to become pregnant; intention to become pregnant again).^19^ These phases are each characterised by specific motivations and receptiveness and thereby highlight the need for targeted intervention approaches at each phase. In line with this, our findings suggest that, in addition to general public health initiatives, efforts focused on the needs of people who may become pregnant or a parent are also required (dual strategy). These interventions and services should be co-developed with patients and the public to ensure they are acceptable, appropriate, and address structural, cultural and gender-related barriers.

Recommendations for policy and clinical practice to tackle inequalities in preconception health and care reflect the need for targeted approaches at interpersonal, institutional, and environmental/societal levels. Previous calls for action have mainly focussed on recommendations at institutional and environmental/societal levels, including calls for preconception health education in schools and integration of (incentivised) preconception care within existing services that are accessible to all.^20-22^ In addition, workshop participants also defined recommendations at the interpersonal level, highlighting the need to involve members of the community to raise awareness and provide peer-support, to normalise respectful and inclusive conversations about pregnancy planning and preparation in diverse local communities. No recommendations were identified at the intrapersonal level, which supports calls to move away from a focus on individual responsibility which often evokes blame and may worsen inequalities.^23^

To our knowledge, recommendations to tackle inequalities in preconception health and care have not previously been co-developed by bringing together multi-disciplinary professionals as well as people with lived experience. Our workshop findings were further strengthened by the hybrid (in person and online) format, and involvement of professionals at all career stages, which allowed for a wide range of views and experiences to contribute to the discussions and recommendations. While previous calls for action have been developed by researchers and policy makers,^20-22^ the inclusion of people with lived experience in our workshop may have identified additional recommendations, in particular at the interpersonal level. Our workshop was, however, mainly attended by UK participants. Discussions were therefore largely focussed on the UK context of inequalities and preconception intervention gaps, and barriers and opportunities not discussed during our workshop may exist in other countries. In the discussion on recommendations, workshop participants noted that we can learn from best practice models in other countries. This may include approaches to pregnancy intention screening being tested in countries such as the USA and Sweden,^24, 25^ and the national ‘Solid Start’ programme in the Netherlands.^26^ Lastly, mandatory folic acid fortification to prevent neural tube defects was not discussed as part of the workshop recommendations. This may be because it was used in the presentation at the start of the workshop to highlight an exemplary public health policy to reduce inequalities.

Inequalities in preconception health and care are rife and can have a detrimental impact on the health and wellbeing of current and future generations. Our co-developed recommendations provide a guiding framework to address and reduce inequalities at interpersonal, institutional, and environmental/societal levels. Further research that highlights the scope of preconception health and care inequalities and structural barriers, and produces co-developed and evidence-based interventions, will further strengthen advocacy for implementation of the recommendations.

## Data Availability

All data produced in the present work are contained in the manuscript.

## Author contributions

DS, JH, CS, SJH, EHC, MB, AAWJ, MC and SC developed the idea and agenda for the workshop. DS and JH co-chaired the workshop. DS, CS and SC summarised the findings, and DS and SC wrote the first draft of the manuscript. JH, CS, SJH, EHC, MB, AAWJ and MC reviewed and edited the manuscript draft. All UK Preconception EMCR Network conference workshop participants (group members listed below) contributed to the workshop discussions and co-development of the recommendations, and reviewed, edited (optional) and approved the final manuscript.

## UK Preconception EMCR Network conference workshop participants (who agreed to be listed as co-authors)

Alkis Velivasis, Amy Hough, Anita Banerjee, Bernadette Jenner, Caitlin Victoria Gardiner, Chamendi Nishshanka, Cheryl McQuire, Dorothea Geddes-Barton, Eleanor Dyer, Eleonora Hristova-Atanasova, Elpida Vounzoulaki, Emma Brough, Evita Pappa, Fiona Ghalustians, Hannah Webb, Hope Jones, Jayne Walker, Kate Nash, Khadija Choudhury, Lucy Goddard, Majel McGranahan, Manjiri Khare, Maria Memtsa, Merissa Elizabeth Hickman, Michael Daly, Nathan Davies, Nicola Vousden, Niharika Manu, Noreth Muller-Kluits, Pauline Cross, Rhea Khosla, Rita Forde, Ruth Kipping, Shivali Lakhani, Stacey Draper, Tamara Wieles, Tsakani Hlungwani, Yves Jacquemyn.

